# Psychological Distress Among People Losing Work During the COVID-19 Pandemic in Australia

**DOI:** 10.1101/2020.05.06.20093773

**Authors:** Alex Collie, Luke Sheehan, Caryn van Vreden, Genevieve Grant, Peter Whiteford, Dennis Petrie, Malcolm R Sim

## Abstract

**Introduction:** This study estimated the extent of psychological distress among people losing work during the coronavirus disease of 2019 (COVID-19) pandemic in Australia, and examined associations between distress, nature of work loss and degree of social interaction.

**Methods:** Data were from a baseline online survey of an inception cohort recruited in the weeks following the introduction of physical distancing and movement restrictions to contain the spread of COVID-19 in Australia. These restrictions resulted in widespread unemployment and working hour reduction. Psychological distress was measured using the Kessler-6 scale. Data on nature of work loss, social interactions, demographic, job and occupational characteristics were also collected. Regression modelling was conducted to determine the relationship between work loss, social interactions and psychological distress, accounting for confounders.

**Results:** Among the 551 study participants 31% reported severe psychological distress, 35% in those with job loss and 28% in those still employed but working less. Those who had significantly greater odds of high psychological distress were younger, female, had lost their job and had lower social interactions. The relationship between job loss and distress became non-significant when financial stress, and occupation were included in the regression model, but the protective effect of higher social interactions remained significant.

**Discussion:** There was a high prevalence of psychological distress in people losing work during the coronavirus pandemic. Age, gender, job loss and social interactions were strongly associated with distress. Interventions that promote social interaction may help to reduce distress during among people losing work during the COVID-19 pandemic.

## Introduction

In response to the coronavirus pandemic governments internationally have sought to contain viral spread by limiting travel and physical interaction between citizens. These measures have led to large-scale unemployment. In the USA 26 million people filed applications for unemployment insurance benefits in the four weeks to 24^th^ April 2020 (1). In Australia an estimated 800,000 people have lost their jobs in the weeks after social distancing measures were introduced with unemployment expected to more than double (2).

The harmful impacts of unemployment on mental health are well described (3), including in people whose work is impacted by viral epidemics (4, 5). These effects may be mitigated by social connections (6). The protective effect of social networks may be less during the coronavirus pandemic as physical distancing presents a barrier to social interaction. Policy responses to the coronavirus pandemic thus introduce the potential for a psychological ‘double whammy’ of job loss combined with reductions in social interaction.

This study aimed to characterise the prevalence of psychological distress in people losing work or losing their jobs during the pandemic, and to determine the associations with social interactions. We hypothesised that job loss and lower levels of social interaction would increase the likelihood of psychological distress.

## Methods

### Study design and procedures

We report findings from the baseline data collection of an inception cohort study of people losing work, or losing their jobs, during the COVID-19 pandemic. Participants were recruited through social and mainstream media. Inclusion criteria included age 18 years or over, working in a paid job during the final three months of 2019, and loss of job or reduction in working hours during the COVID-19 pandemic beginning early 2020. All participants included in this paper were enrolled and completed the baseline online survey between 27^th^ March and 20^th^ April 2020.

The study was approved by the Monash University Human Research Ethics Committee (Project ID 24003).

### Outcome

Psychological distress was assessed using the Kessler 6 questionnaire (7). The score was calculated and categorised into low, moderate and severe psychological distress as per established criteria (8, 9).

### Exposure Variables

Work status was dichotomised as having lost employment completely (job loss) or employed but working less.

Social interactions were measured using the Social Interaction sub-scale of the Duke Social Support Index (DSSI) (10). The scale ranges from 4 (least social interaction) to 12 (most social interaction).

### Other Variables

Financial stress was assessed with the question ‘If all of a sudden you had to get $2000 for something important, could the money be obtained within a week? (11) Response options were ‘yes’, ‘no’, and ‘don’t know’. For analysis ‘don’t know’ and ‘no’ were collapsed.

Participants selected the Australian and New Zealand Standard Classification of Occupations major group that best fitted their occupation. The categories ‘Technicians and Trade Workers’, ‘Machinery Operators and Drivers’, and ‘Labourers’ were combined into one group for analysis due to small sample size.

### Data Analysis

Frequencies and percentages were used to describe the sample given most outcomes were categorical. Mean and standard deviation was used for the social interaction as this variable is continuous. The effect of job loss and social support on psychological distress was modelled using ordinal logistic regression. Two models were estimated. The first contained only the two explanatory variables of interest: work status and social interaction along with age and gender. The second, adjusted model was based on the first but with confounder variables added. A variable was included in the model if it altered the coefficient for work status or social interaction score by more than 10% when added to the first model.

## Results

551 participants were included in analysis, and had no missing data on all variables required for the regression models. The sample was over three quarters female and 61.2% were aged between 45 and 65 years (Table 1). Approximately one third (31.0%) were experiencing severe psychological distress and a further 45.9% were experiencing moderate distress. Almost two-thirds had lost their job. The percentage experiencing severe distress was greater in the job loss group (35.2%) than in the still employed group (28.1%).

**Table 1.**
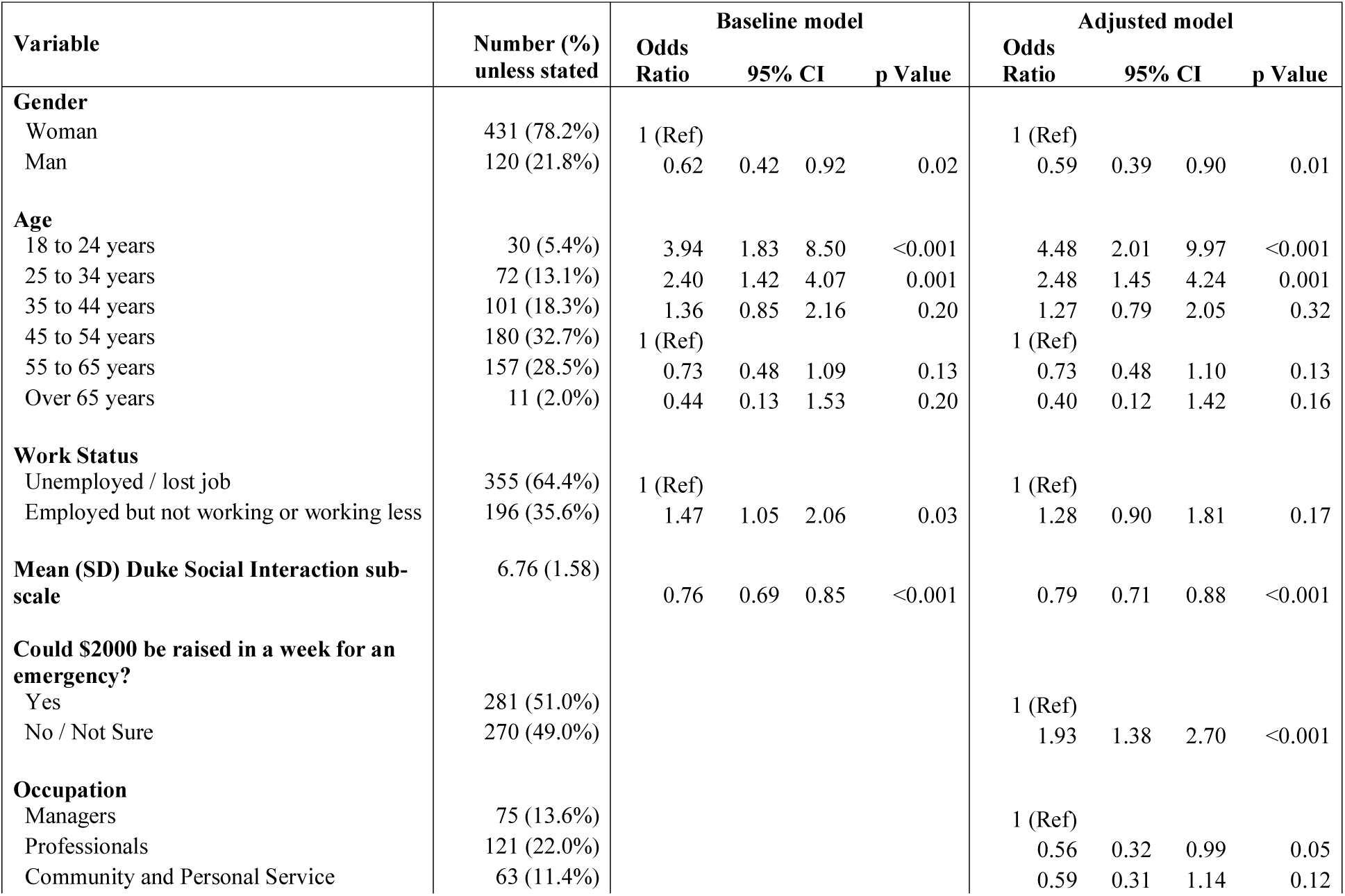
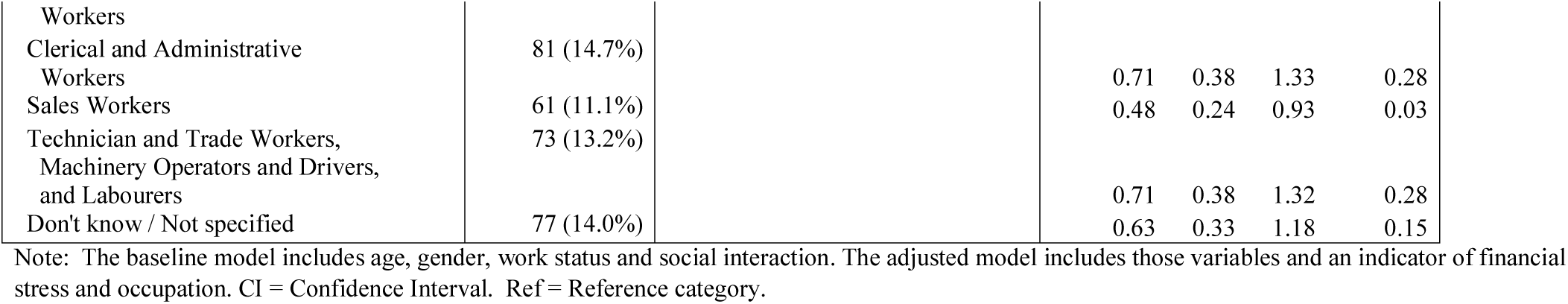
Results of the ordinal logistic regression models with psychological distress as the outcome.

### Baseline Model

Both work status and social interaction had a significant effect on psychological distress in the baseline model. Each 1-point increase in the DSSI score was associated with 0.76 (95% CI: 0.69-0.85) reduction in the odds of psychological distress. Participants who had lost their job had greater odds of reporting psychological distress [OR=1.47, 95% CI: 1.05 – 2.06]. Women and those aged under 35 years also had greater odds of distress in this model.

### Confounders

Financial stress and occupation confounded the interaction between outcome and exposure variables and were included in the adjusted model. A range of other variables were tested but did not alter this interaction including: being the primary earner in the household, employment type prior to changes (full-time/part-time/casual), whether an application for social assistance had been filed, self-assessed health, highest education level, job tenure, job satisfaction, days of notice before change in job status, industry, and household structure.

### Adjusted Model

The addition of the two confounders altered the estimated effect of the exposure variables on psychological distress. The effect on the DSSI score was minimal, with the estimated odds ratio for a 1-point increase in the DSSI score changing to 0.79 (95% CI: 0.71, 0.88). There was a larger attenuation on the effect of work status and this effect was no longer significant (OR = 1.28, 95% CI: 0.90, 1.81). As in the baseline model, females and younger workers had greater odds of psychological distress. Those in financial stress had elevated odds of psychological distress (OR: 1.93, 95% CI: 1.38, 2.70). Managers were the occupation with the largest odds of experiencing psychological distress, with both professionals and sales workers having approximately only half the odds of distress.

## Discussion

This study of Australians losing work or losing their jobs during the COVID-19 pandemic found that 76% of participants reported moderate or severe psychological distress. The proportion with severe psychological distress (31%) is approximately four times that observed in employed Australians aged 18 to 65 years (~8%) and more than 2.5 times the rate observed in Australian adults (12). These findings extend recent Australian government data showing a higher than normal prevalence of stress, anxiety and hopelessness among the general community during the COVID-19 pandemic (2)

A greater proportion of people experiencing job loss reported severe psychological distress than those still employed. This difference was statically significant in a baseline model adjusting for age, gender and social interaction, but non-significant when other confounders were included in an adjusted model. This finding suggests that the pathway from job loss to psychological distress may be mediated by financial stress and occupation. In contrast the effect of social interactions remained robust and statistically significant in both baseline and adjusted models. People reporting greater social interaction were less likely to report psychological distress. Maintaining or enhancing social interactions may be a strategy that can support positive psychological outcomes in people experiencing employment shocks during the COVID-19 pandemic.

Study limitations include its cross-sectional nature and reliance on self-report. The sample may not be representative of the population affected. Strengths include the use of validated measurement instruments, the temporal proximity of data collection to job and work loss and that the analysis accounted for multiple confounders.

To our knowledge, this is one of the first studies examining psychological distress specifically among people losing work during the COVID-19 pandemic. Many nations are experiencing substantial economic downturns and sharp rises in unemployment. Further studies of psychological complaints amongst people whose work has been adversely impacted during the COVID-19 pandemic, and strategies to mitigate any negative impacts, are warranted.

## Data Availability

De-identified study data is available by contacting the first-named author.

## Notes

### Competing Interest Statement

The authors have declared no competing interest.

### Funding Statement

At the time of submission this study had received financial support from Monash University.

